# Assessment of correlations between risk factors and symptom presentation among defined at-risk groups following a confirmed COVID-19 diagnosis

**DOI:** 10.1101/2021.11.30.21267029

**Authors:** Dylan Aidlen, Jamie Henzy

**Affiliations:** Department of Biology, Northeastern University, Boston, Massachusetts, USA

**Keywords:** symptom presentation, autoimmune, at-risk groups, symptom severity

## Abstract

This study analyzes the specific linkages between symptoms within individual COVID patients belonging to at-risk groups. The goal was to determine how strongly linked patient symptoms are within these at-risk groups to find any associations between factors such as comorbidities and COVID symptoms. In this study, de-identified patient data from the N3C database was utilized in order to link representative immunocompromised states with specific symptoms, and non-immunocompromised state with the same, to determine if the strength of the correlation changes for these at-risk groups. Multiple autoimmune disorders resulting in immunocompromised state were analyzed, to determine if severity of immune response and inflammatory action plays a role in any potential differences. An exploratory approach using statistical methods and visualization techniques appropriate to multidimensional data sets was taken. The identified correlations may allow pattern analysis in disease presentation specific to a given population, potentially informing pattern recognition, symptom presentation, and treatment approaches in patients with immune comorbidities.

## Introduction

Toward the end of 2019, a novel coronavirus in Wuhan led to a group of pneumonia cases, following which it rapidly spread^(1)^. This resulted in an epidemic throughout China, after which it spread across the world, leading to a large number of cases. The World Health Organization termed the novel coronavirus COVID-19^(2)^.

COVID-19 is caused by SARS-CoV-2, severe acute respiratory syndrome coronavirus 2, and it is a coronavirus that has had a significant impact. More than 184 million cases have been confirmed as of this point, with approximately 4 million deaths. COVID-19 is characterized by its rapid spread and by its notable variety in clinical presentation and symptom severity. A large number of those afflicted remain asymptomatic, with current estimates ranging from 33 percent of those infected with SARS-CoV-2 up to 77 percent^(3, 4, 5, 6, 7, 8)^. It has also been proposed that some of those who are recorded as asymptomatic may be presenting with unnoticed clinical abnormalities ^(9, 10)^. Regarding COVID positive patients who are symptomatic, mild disease was found in roughly 80 percent, severe disease in roughly 15 percent, and critical disease in roughly 5 percent. The overall fatality rate is currently reported at roughly 2 percent^(11,12,13,14,15,16,17,18,19)^. The fatality rate among hospitalized COVID positive patients is estimated to fall somewhere in the range of 10 to 20 percent^(20)^.

This study aims to look specifically at COVID positive patients with type 1 diabetes, vitiligo, and alopecia. Alopecia is an autoimmune disorder targeting hair follicles and vitiligo is an autoimmune/inflammatory disease that attacks pigment cells/appears to focus on the epidermal layer ^(21,22)^. In contrast to these two disorders, type 1 diabetes is focused on the pancreas/insulin, and studies have tied COVID to elevated blood levels of amylase and lipase, including acidosis, renal failure and diabetes ^(23,24)^. This study analyzes the specific linkages between symptoms within individual COVID patients belonging to these at risk groups and first compares them to the overall group of COVID positive patients. Following this comparison, this study compares symptom presentation and severity within and between these at-risk groups.

The goal of this study was to utilize this set of immune disorders that vary in their severity and location of impact to see if there is a pattern in symptom presentation and severity that may be preserved across these immune groups, and which significantly differs from the symptom presentation of the overall population. This study has the potential to begin elucidating any differences in symptom presentation in the immune groups, with the possibility of generalizing the results in the future if there are presentation patterns that are well preserved across the immune groups tested. A secondary goal was to determine if there is the potential for autoimmunity/inflammatory action alone to significantly impact COVID-19 presentation and severity, or if the severity and area impacted by the autoimmune disorder was vital. This will also help to delineate whether the inflammatory action/autoimmunity itself was responsible for the majority of any potential differences from the controls, or if severity/correlation to areas COVID impacts the most would play a large role. These three immune groups represent a range of autoimmune/inflammatory responses and had a large cohort size.

## Methods

### Overview

This project was carried out in two phases. Phase one involved determining if there is a significant difference between a symptom’s occurrence when compared with occurrence of every other symptom studied in COVID-19 patients within the overall group and within each comorbid autoimmune disorder group. Phase two involved determining is there is a significant difference in symptom presentation in COVID-19 patients with comorbid autoimmune disorders of differing severity and impacted location.

The analysis involved three experimental groups: type 1 diabetes, vitiligo, and alopecia patients who were COVID positive. The control groups involved the group of all patients who were COVID positive.

We utilized Slate, Contour, and Workshop, the N3C NIH Collaborative Portal’s built-in tools for manipulating and analyzing data in order to isolate these experimental and control groups, and then make the comparisons. The following step was to determine if there was a significant difference between symptoms in a typical COVID-19 patient/one without comorbidities in comparison to a COVID-19 patient with one of the three autoimmune comorbidities of varying severity being analyzed. Following this, symptom presentation was compared between the three experimental groups of autoimmune disorders of varying severity in order to see if there was a significant difference in presentation and outcome between these at-risk groups.

### Data Origin

The data utilized comes from the N3C Enclave, which is an endeavor led by the National Center for Advancing Translational Science (NCATS) and is a centralized, harmonized, high-granularity EMR repository ^(25)^. Methods utilized for data collection, database design, and data harmonization have been detailed prior ^(26)^. The Observational Medical Outcomes Partnership (OMOP) is utilized as the primary data model that enables harmonization, and each study area provides information to the central data repository using the standard OMOP definitions and terms or is converted to meet OMOP standards.

### Creation of concept sets

We created concept sets using Atlas ^(27)^. For each of the chosen symptoms, we found all records, codes, IDs, and domains relating them across all classification vocabularies (SNOMED, RxNorm, Nebraska Lexicon, etc). We then looked at the full hierarchy of each potential term for the symptom, including all descendants and excluding any descendants who did not fit the symptom criteria, or whose symptom presentation did not match what is expected of COVID. Concept sets were generated and utilized for fourteen of the primary symptoms of COVID: Breathing abnormalities, fever or chills, congestion or runny nose, fatigue, sore throat, chest pain or pressure, impairment of consciousness, diarrhea, impairment of taste or smell, cough, headache, nausea or vomiting, body/muscle pain and aches, and rash; these concepts incorporate the group of potential terms and IDs that could refer to each of these symptoms (Supplementary Table 1). After generating a concept set of all potential IDs/codes, we then created and generated a new concept set with these specifications in the N3C Enclave. The three immune groups of Alopecia, Type 1 Diabetes, and Vitiligo were also matched to the relevant Atlas IDs and a concept set was generated for each.

#### Defining COVID positive and symptom occurrence

We filtered the full de-identified dataset for COVID positivity using as criteria a positive Lab measurement (PCR or AG), positive Antibodies, or Positive Covid Diagnosis (corresponding to U07.1 ICD10-CM code). Once filtered, we were left with 2,182,104 positive patients. We define symptom occurrence as an individual who possesses at least one symptom definition that falls within the chosen concept set, where the onset of that symptom was within +/-one month of the COVID infection date. We then went through all of the patients for all of our condition concepts and experimental groups (alopecia, vitiligo, and diabetes), and created a 2,182,104 by 18 table. Column 1 included patient id, columns 2-15 included each symptom (along with a 1 if present, and a 0 if not), and then our three experimental groups (also with a 1 or 0). This involved utilization of the N3C Enclave’s built-in data analysis tools, as data cannot leave the Enclave. We utilized Contour, Slate, Fusion, Quibit, and Quiver for preliminary analyses, and pySpark and the code workbook/code repository were utilized for additional analysis.

#### Statistical Analysis

We looked at patients in four groups: all COVID positive patients (2,182,104 positive patients), COVID positive patients with alopecia (1,864 patients), COVID positive patients with vitiligo (2,004 patients), and COVID positive patients with type 1 diabetes (10,265 patients). First, percentages of each symptom for each group were generated (Fig1a-Fig1d). Then we applied the Chi-square test of independence (for categorical variables) to see if there was a statistically significant difference in occurrence for each of the 14 symptoms across the four study groups (Fig 2). We also applied the Chi-square test of Independence to compare each symptom within each group, with a 14×14 table for each group of the result in order to analyze potential relationships between variables. Heatmaps were generated to depict this, displaying the p-values for all four COVID positive groups (Fig 2-5). We depicted the p-value heatmaps with increasing intensity as value decreases.

**Fig 1a:**
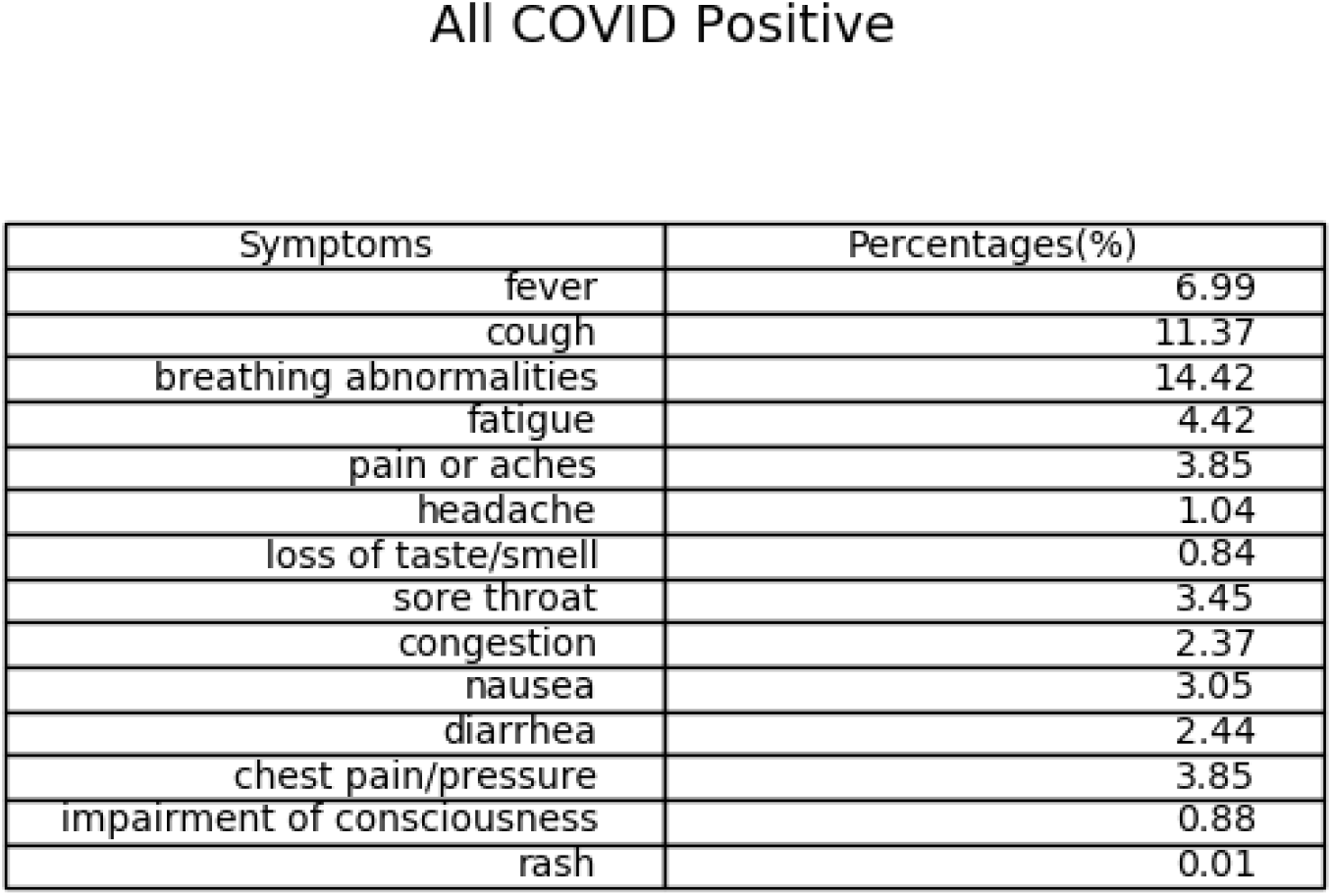
Percentage table for population of all patients who are COVID positive

**Fig 1b:**
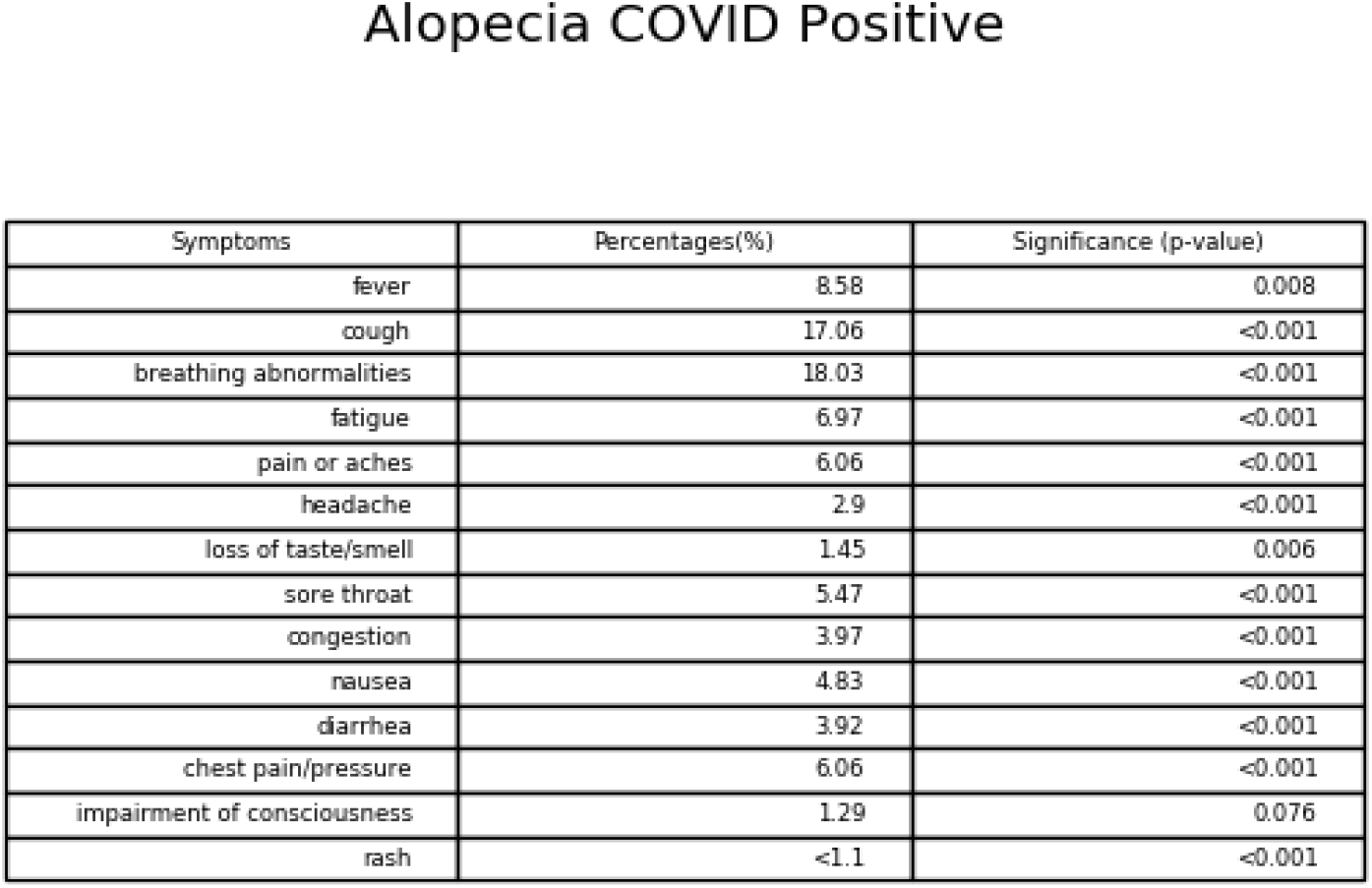
Percentage table for population of all patients who are COVID positive and have Alopecia. Significance between Alopecia and each symptom is included

**Fig 1c:**
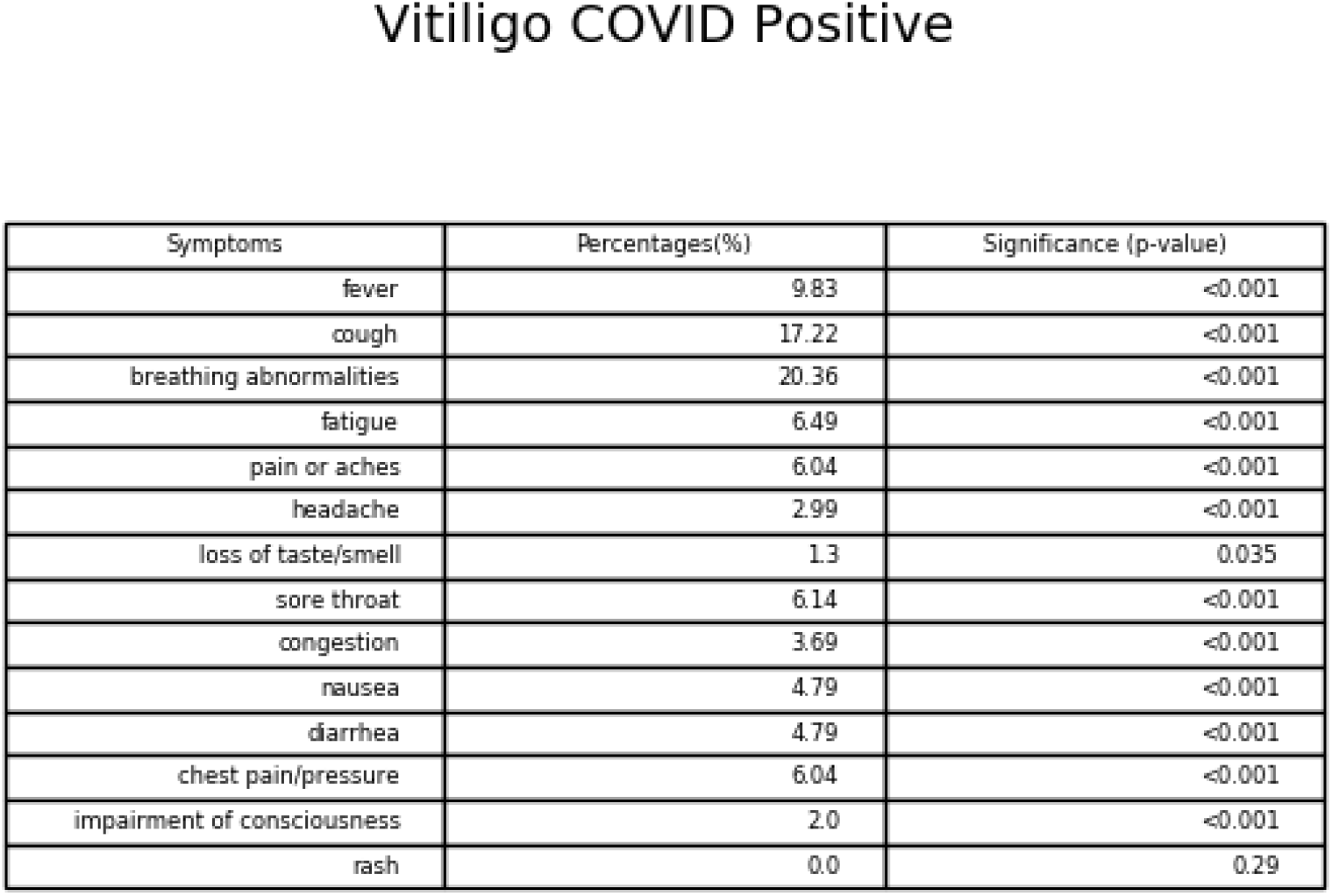
Percentage table for population of all patients who are COVID positive and have Vitiligo. Significance between Vitiligo and each symptom is included

**Fig 1d:**
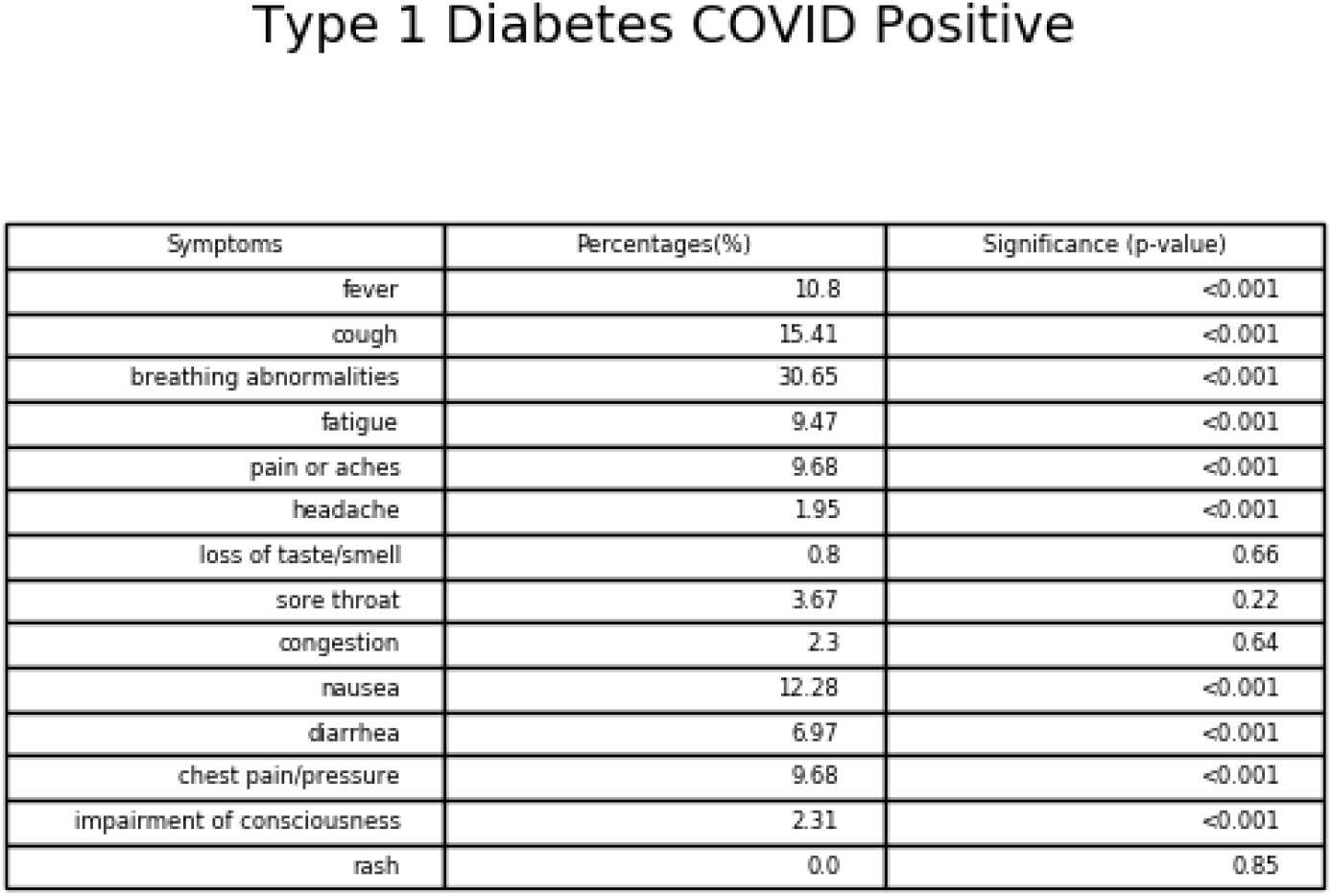
Percentage table for population of all patients who are COVID positive and have Type 1 Diabetes. Significance between Type 1 Diabetes and each symptom is included

**Fig 2:**
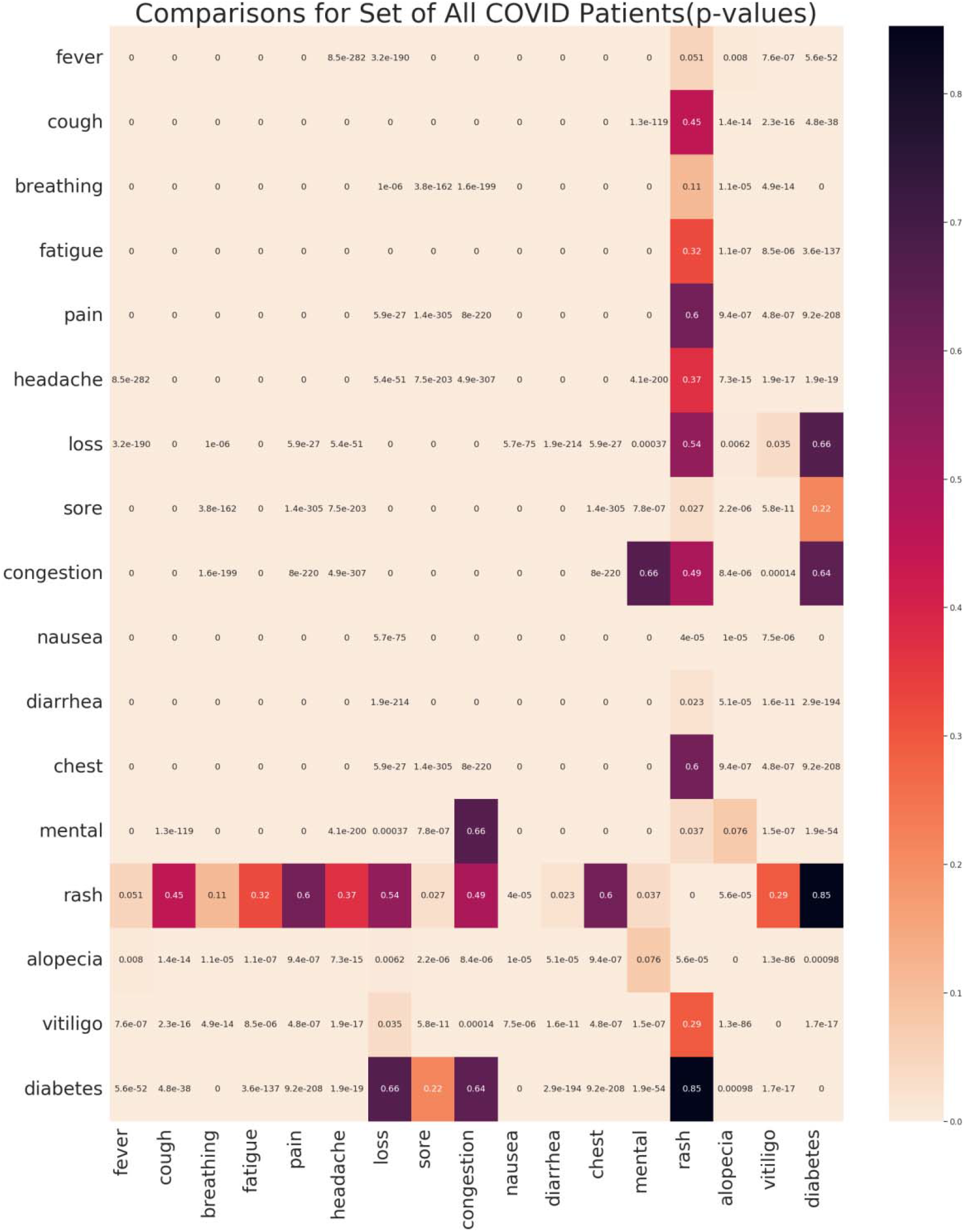
Heatmap of p-values in set of All COVID positive patients for comparison of all symptoms to every other symptom and comparison to each of the three immune groups (alopecia, vitiligo, and type 1 diabetes); color intensity increases with decreasing value; the following abbreviations were utilized: breathing (breathing abnormalities), loss (loss of taste/smell), chest (chest pain/pressure), mental (impairment of consciousness)

## Results

### COVID-19 Symptom Percentages among all groups

We observed higher percentages across all immune groups for all symptom categories, except for Type 1 Diabetes in the loss of taste/smell and congestion categories, which contained percentages similar but slightly lower than those seen in the overall group of COVID positive patients. We also observed approximately 0% of patients falling into the rash group except for the group of alopecia patients, in which <1% of patients developed a rash within +/-one month of COVID onset.

### COVID-19 Significance Results in relation to immune comorbidity

We observe a significant relationship between all three immune groups and the group of fourteen symptoms, except for a few exceptions. For the alopecia immune group, the exceptions are a lack of significance for the mental (impairment of consciousness) symptom group. For the vitiligo group, the exceptions are a lack of significance for the rash group; this is expected, as the vitiligo patient percentage with rash as a symptom was 0%. For type 1 diabetes, the exceptions are a lack of significance for the rash group (also 0% in this immune group), congestion, sore throat, and loss of smell/taste.

### COVID-19 Significance Results in relation to symptom comparisons

In the overall COVID positive group, we observe significance between symptoms for most symptoms, with a few exceptions. We failed to reject the null hypothesis for the relationship between rash with all symptoms except for sore throat, nausea, diarrhea, and mental symptoms. We also were not able to conclude a significant relationship for mental symptoms and congestion symptoms.

In the alopecia COVID positive group, we observed drastically more interactions that we failed to reject the null hypothesis for. For the fever symptom, we failed to conclude significance when looking at its interactions with pain, headache, chest pain, mental symptoms, and rash. For cough, the interaction with loss of smell/taste, mental symptoms, and rash were all not significant. For breathing abnormalities, headache, loss of smell/taste, and rash were all not significant. Significance of additional interactions can be observed in Fig3b for the alopecia group, Fig4b for the vitiligo group, and Fig5b for the type 1 diabetes group. Of note, all three immune groups experienced many more instances where significance could not be concluded, and associations between symptoms could not be determined. We observed that the symptom interactions where we failed to reject the null hypothesis occurred between roughly the same symptoms in all three immune groups, while these symptom interactions had significant relationships in the overall set of all COVID positive patients.

**Fig 3:**
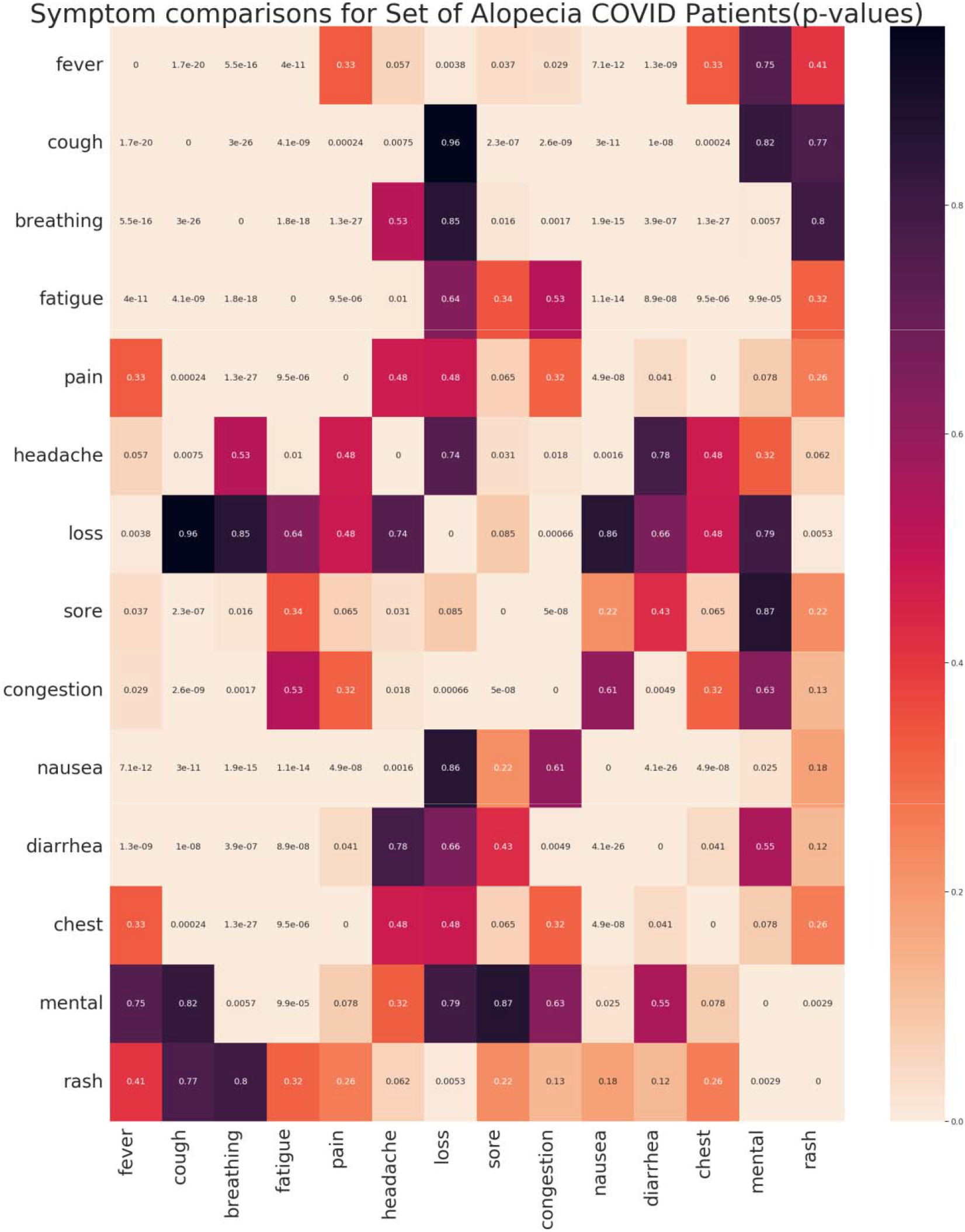
Heatmap of p-values in set of Alopecia COVID positive patients for comparison of all symptoms to every other symptom; color intensity increases with decreasing value

**Fig 4:**
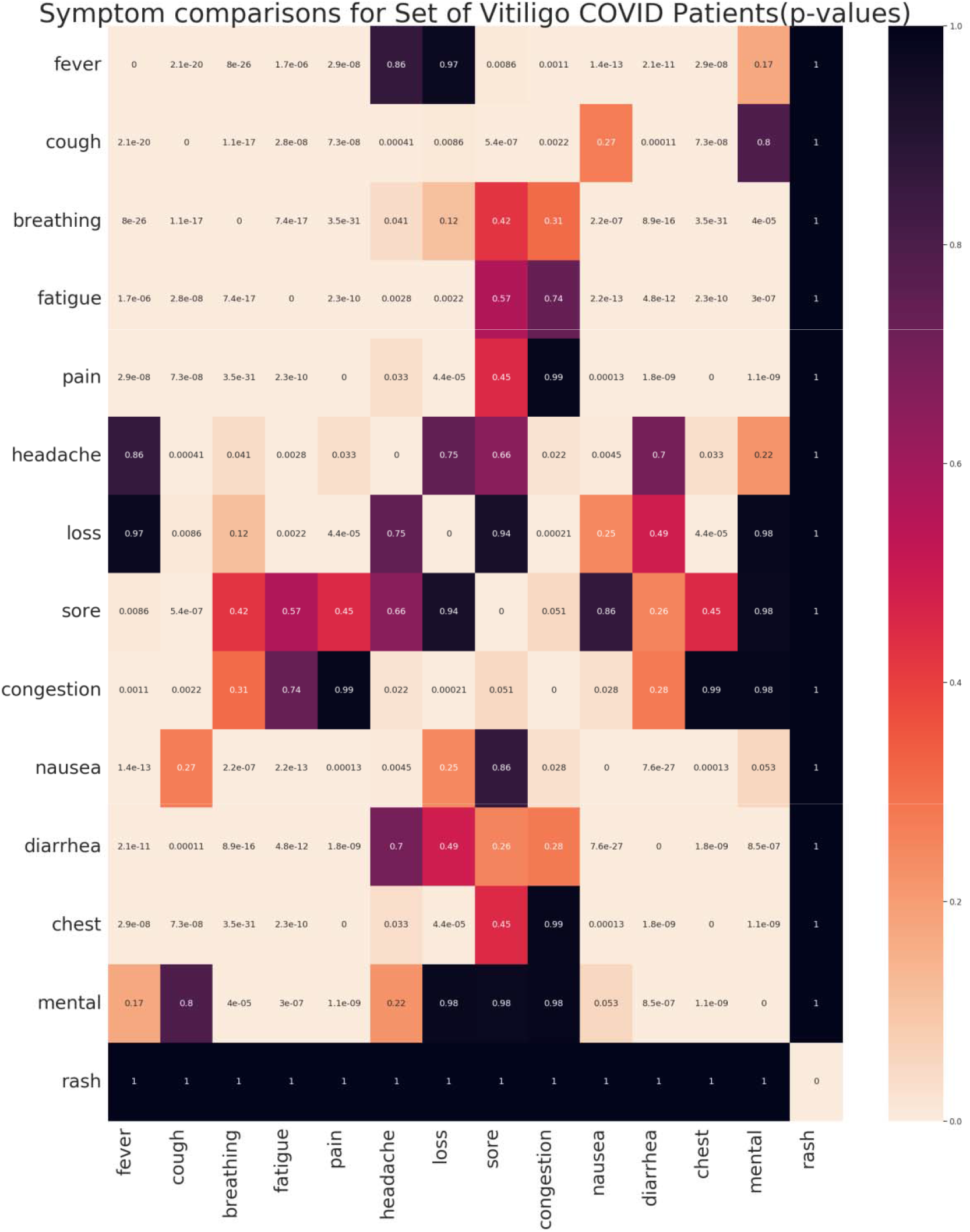
Heatmap of p-values in set of Vitiligo COVID positive patients for comparison of all symptoms to every other symptom; color intensity increases with decreasing value

**Fig 5:**
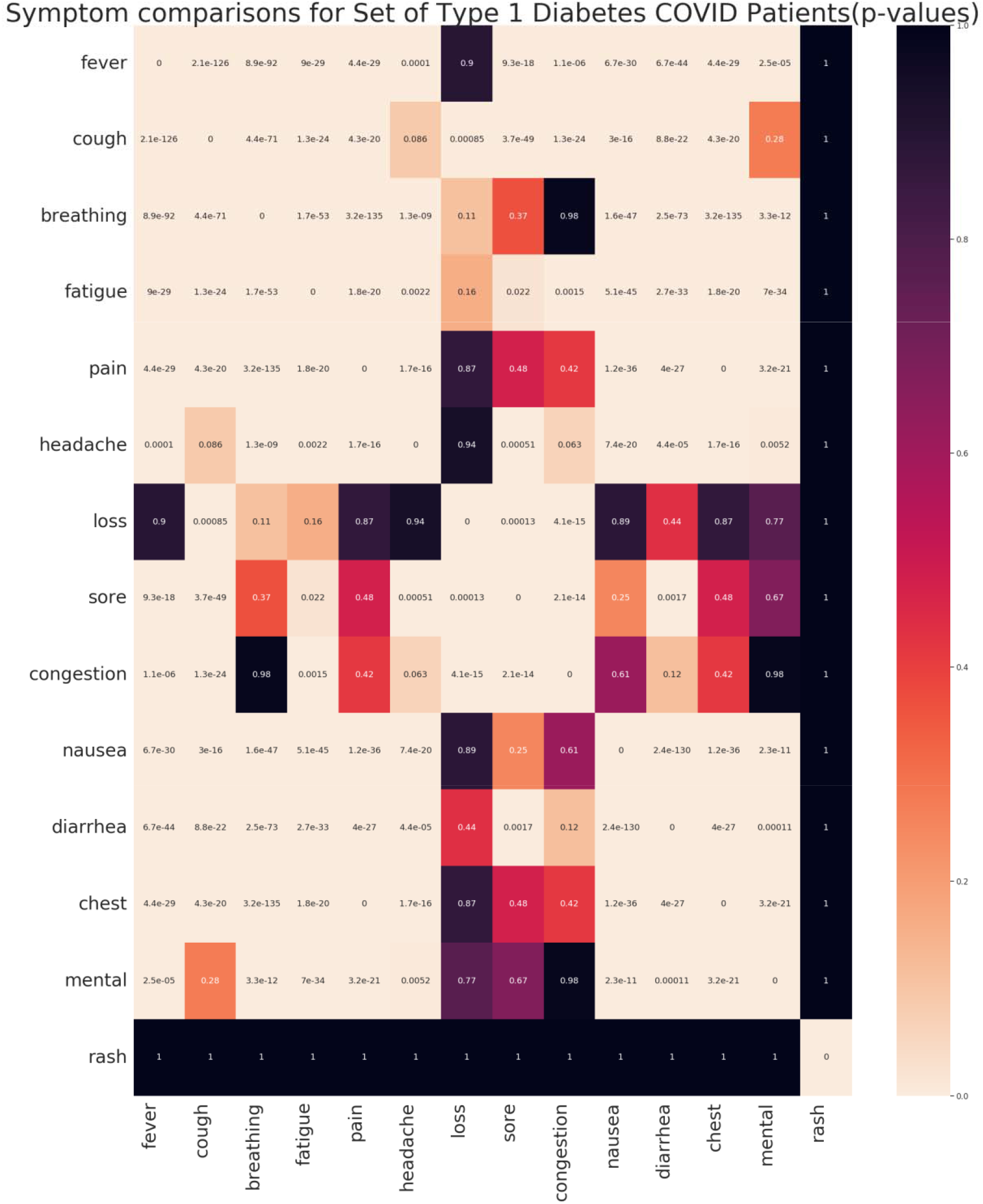
Heatmap of p-values in set of Type 1 Diabetes COVID positive patients for comparison of all symptoms to every other symptom; color intensity increases with decreasing value

## Discussion

Our analysis indicates that there is a significant difference in symptom presentation between the overall population of COVID-19 positive patients and the COVID positive patients with the three tested immune comorbidities. We observe a distinct but preserved symptom presentation pattern across all three immune groups, and our results do not appear to indicate that the severity of the autoimmune disorder impacts symptom occurrence. Interestingly, our results show that while symptoms are significantly associated in almost every interaction in the overall set of COVID positive patients, many symptoms in the immune groups do not experience this significant association. This means that while symptoms are likely correlated with immune disease itself, symptoms experienced by patients within those immune groups are less likely to be correlated. The overall population of COVID positive patients experienced much more symptom to symptom correlations than the immune groups did.

This data reveals that diagnostic approaches and model creation regarding immune patients with COVID-19 might require extra care and different inputs. The results of this study reveal potential symptoms researchers should be focusing on when studying immune patients with COVID-19. This differing symptom presentation that is preserved across all three immune groups may serve as a potential aid in more tailored diagnostic tools and modeling approaches for immune co-morbid COVID-19 patients.

Patients in the immune group also experienced higher percentages of symptoms for almost every symptom, though in this case there did not appear to be a difference in severity of percentages across immune groups. Cough and breathing abnormalities within +/-30 days of COVID diagnosis was also particularly prevalent in all three immune groups when compared to the overall set of all COVID positive patients.

There have been many studies examining the potential role of autoimmune disorders in severe COVID-19. The results have been conflicting as to whether autoimmune disorders increase susceptibility and likelihood of COVID-19, and their role in severity ^(28, 29, 30, 31, 32)^. Prior to this study, there has been a lack of studies that consider symptomology differences in addition to severity in general or susceptibility. From the results of this study, it is evident that autoimmune disorders impact symptomology, presentation, and potentially severity of COVID-19 disorder.

There are some recent studies examining emergent autoimmune symptoms following COVID-19 infection, with COVID-19 being suspected as the cause ^(42, 44)^. Given the results of this study, it is possible that existing symptoms of the chosen autoimmune disorders are impacting the interaction between COVID-19 symptoms, which could explain this lack of significance ^(37, 38, 39 40)^. This would require further study, potentially by looking at symptoms reported prior to COVID infection and post COVID infection in these groups. Regardless of the cause, this impact on symptomology will likely impact the speed with which accurate diagnosis of COVID can be made in these groups.

Some recent studies have also claimed that COVID-19 can lead to the emergence of new autoimmune disorders following COVID infection^(33, 34, 39, 40, 42, 44)^. Based on the results from this study, this claim could be assessed by analyzing this group of people in comparison to the groups from this study and the overall group, to see whether their symptoms correspond more closely to the general population symptoms, or to the symptoms that appear to be shared across all three immune groups. For example, since the autoimmune disorders in this study do show significantly differing symptom presentation, if those who “develop” autoimmune disorders following COVID-19 display significant overlaps with these symptoms, it is possible that they had an undiagnosed immune disorder, or that COVID-19 exacerbated an existing autoimmune issue. If the symptoms experienced during COVID infection do not match between these two groups, it might be more likely that some action of COVID-19 itself more directly led to this autoimmune presentation.

In conclusion, patients across all three immune groups were more likely to be symptomatic than patients in the overall population of COVID positive, across nearly every symptom, for all three immune groups. There was also no significant variation or severity between the three immune groups when it came to symptomology or severity. There also appeared to be fewer associations between symptoms in all three of the immune groups, with all three immune groups experiencing this lack of association with a similar distribution. These results call for more care to be taken when generating models and building diagnostic tools for COVID in people with preexisting immune conditions. They also call for a more targeted COVID assessment and treatment process for patients with immune comorbidities which give extra care to symptoms that maintain their associations even in the immune groups as determined by this study.

## Conclusion

This study determined there is a significant difference between symptom presentation in COVID-19 patients with a comorbid autoimmune disorder when compared to the general population of COVID positive patients. Patients across all three immune groups were more likely to be symptomatic than patients in the overall population of COVID positive, across nearly every symptom, for all three immune groups. There also appeared to be fewer associations between symptoms in all three of the immune groups, with all three immune groups experiencing this lack of association with a similar distribution. There was also no significant difference in symptom occurrence and severity in COVID-19 patients with comorbid autoimmune disorders of differing severity and impacted location. While all groups appeared to have higher occurrence of symptoms than the general population and appeared to have significant relationships with most symptoms, the difference between immune groups was not found to be significant. This study begins to delineate potential differences in symptomology and associations between symptoms within immune comorbid COVID patients when compared to the overall COVID positive population. The next steps are to note this symptom presentation pattern that occurred across all three immune groups and see if it is preserved across a wider range of immune groups.

## Data Availability

The analyses described in this publication were conducted with data or tools accessed through the NCATS N3C Data Enclave https://covid.cd2h.org and N3C Attribution & Publication Policy v 1.2-2020-08-25b supported by NCATS U24 TR002306. This research was possible because of the patients whose information is included within the data and the organizations and scientists who have contributed to the on-going development of this community resource https://doi.org/10.1093/jamia/ocaa196/5893482.

## Acknowledgements

Authorship was determined using ICMJE recommendations.

The N3C data transfer to NCATS is performed under a Johns Hopkins University Reliance Protocol # IRB00249128 or individual site agreements with NIH. The N3C Data Enclave is managed under the authority of the NIH; information can be found at https://ncats.nih.gov/n3c/resources.

The analyses described in publication were conducted with data or tools accessed through the NCATS N3C Data Enclave https://covid.cd2h.org and N3C Attribution & Publication Policy v 1.2-2020-08-25b supported by NCATS U24 TR002306. This research was possible because of the patients whose information is included within the data and the organizations and scientists who have contributed to the on-going development of this community resource https://doi.org/10.1093/jamia/ocaa196/5893482.

