## Supplemental Table 1 for "Assessment of correlations between risk factors and symptom presentation among defined at-risk groups following a confirmed COVID-19 diagnosis"

**Supplementary Table 1: Concept Definitions Created:**

This table includes the fourteen symptom sets that were utilized in this study, and all of the relevant terms that are encompassed by the given symptom set.

| **Symptom/Concept Set** | **Concepts within set** |
| --- | --- |
| Breathing abnormalities | Platypnea, Borg Breathlessness Score: 2 slight, Borg Breathlessness Score: 3 moderate, Borg Breathlessness Score: 4 somewhat severe, Borg Breathlessness Score: 5 severe, Borg Breathlessness Score: 7 very severe, Borg Breathlessness Score: 10 maximal, Borg Breathlessness Score: 0.5 very, very slight (just noticeable), Breathless - strenuous exertion, Medical Research Council Dyspnoea scale grade 1, Medical Research Council Dyspnoea scale grade 2, Medical Research Council Dyspnoea scale grade 3, Medical Research Council Dyspnoea scale grade 5, Borg Breathlessness Score finding, Dyspnea, class III, Dyspnea, Finding of respiration, Nocturnal dyspnea, Labored breathing, Cannot blow, Winded, Dyspnea raising arms, Pneumonia, Pleural plaque, Inspiratory dyspnea, Ease of respiration - finding, Abnormal breathing, Acute respiratory distress, Expiratory dyspnea, Increasing breathlessness, Viral pneumonia, Respiratory distress, Dyspnea, class IV, Dyspnea, class II, Acute respiratory failure, Acute respiratory distress syndrome due to disease caused by Severe acute respiratory syndrome coronavirus 2, Acute respiratory distress syndrome, Orthopnea, Paroxysmal nocturnal dyspnea, Dyspnea after eating, Paroxysmal dyspnea, Minimal breathlessness, Chronic dyspnea, Dyspnea on exertion, Disorder of respiratory system, Acute pulmonary edema, Unable to complete a sentence in one breath, Respiratory failure, Borg Breathlessness Score: 0 none at all, Borg Breathlessness Score: 1 very slight, Borg Breathlessness Score: 6 severe (+), Borg Breathlessness Score: 8 very severe (+), Borg Breathlessness Score: 9 very, very severe (almost maximal), Short of breath dressing/undressing, Medical Research Council Dyspnoea scale grade 4, Pneumonia caused by Human coronavirus, Infantile apnea, Respiratory finding, Dyspnea caused by Severe acute respiratory syndrome coronavirus 2, Trepopnea, eMRC (extended Medical Research Council) dyspnoea scale grade 1, eMRC (extended Medical Research Council) dyspnoea scale grade 2, eMRC (extended Medical Research Council) dyspnoea scale grade 3, eMRC (extended Medical Research Council) dyspnoea scale grade 5a, eMRC (extended Medical Research Council) dyspnoea scale grade 4, eMRC (extended Medical Research Council) dyspnoea scale grade 5b, mMRC (modified Medical Research Council) dyspnoea scale grade 0, mMRC (modified Medical Research Council) dyspnoea scale grade 1, mMRC (modified Medical Research Council) dyspnoea scale grade 2, mMRC (modified Medical Research Council) dyspnoea scale grade 3, mMRC (modified Medical Research Council) dyspnoea scale grade 4, Breathless - moderate exertion, Breathless - mild exertion, Dyspnea at rest, Lung function testing abnormal, O/E - dyspnea, O/E - orthopnea, O/E - respiratory distress, Dyspnea, class I, Dyspnea leaning over, Disorder of lung, Difficulty breathing, Gasping for breath |
| Fever or chills | Fever greater than 100.4 Fahrenheit, Swinging fever, Rising phase of fever, Plateau phase of fever, Falling phase of fever, Prolonged fever, Biphasic fever, Fever, diurnal variation, Reversed diurnal fever, Fever defervescence, Central fever, Pattern of fever - finding, Phase of fever - finding, Fever, Low grade pyrexia, Fever of the newborn, Acute rise of fever, Gradual rise of fever, Continuous fever, Staircase fever, Irregular fever, Rapid fall of fever, Gradual fall of fever, O/E - fever, O/E - hyperpyrexia, Fever with chills, Transitory fever of newborn, Pyrexia of unknown origin, Hyperpyrexia, Fever due to infection, Viral fever, Factitious fever, Fever caused by Severe acute respiratory syndrome coronavirus 2, Spiking fever, Cough with fever, O/E - pyrexia of unknown origin, O/E - level of fever, O/E - temperature low, O/E - hyperpyrexia - greater than 40.5 degrees Celsius, O/E - fever - acute rise, O/E - fever - gradual rise, O/E - fever - continuous, O/E - fever - irregular, O/E - fever - fast fall-crisis, O/E - fever-gradual fall-lysis, Sweating fever |
| Congestion or runny nose | O/E - nasal mucosa, Non-allergic rhinitis, C/O nasal congestion, Nasal mucosa moist, Nasal mucosa boggy, Pale nasal mucosa, Swollen nasal mucosa, Blue nasal mucosa, Crusted nasal mucosa, Hyperemic nasal mucosa, Nasal airway patency - finding, Moistness of nasal mucosa - finding, Congestion of mucosa, Respiratory tract congestion and cough, Respiratory tract congestion, Acute rhinosinusitis, Functional congestion, Nasal mucosa dry, Nasal airway patent, Nasal flaring, Nasal symptom, Nasal airway finding, Nasal cavity over-patent, Variation in patency of nasal airway, Alternate nasal obstruction, Problem blowing nose, Blowing nose ineffectual, Nasal mucosa finding, Finding of color of nasal mucosa, Nasal sinus problem, Finding of appearance of nasal mucosa, Congestive non-allergic rhinitis, Hemorrhagic nasal discharge, Rhinitis, Nasal discharge, Catarrhal nasal discharge, Nasal congestion, Purulent rhinitis, Congestion, Vasomotor rhinitis, Congestion of nasal sinus, Active congestion, Foul smelling discharge from nose, Passive congestion, Mucous membrane swelling, Posterior rhinorrhea, Congestion of throat, Nasal mucosa edematous, C/O - catarrh, O/E-nasal discharge-foul smell, O/E - nasal mucosa edematous, O/E - nasal mucosa wet/boggy, O/E - nasal mucosa pale, O/E - nasal mucosa hyperemic, Mouth breathing with nasal obstruction, Abnormal nasal potential difference, Purulent nasal discharge, Acute congestion, Acute passive congestion, Mottled congestion, Nasal obstruction, Disorder of nasal cavity, Non-infective non-allergic rhinitis |
| Fatigue | Tired all the time, Malaise and fatigue, C/O - debility - malaise, Sensation of heaviness in limbs, Exhaustion - physiological, Rapid fatigue of gait, Tires quickly, Tired on least exertion, Quickly exhausted, Exhausted on least exertion, Attacks of weakness, Postexertional fatigue, Postviral fatigue syndrome, Exhaustion, Fatigability, Extreme exhaustion, Fatigue, Occasionally tired, Asthenia due to disease, Heavy legs, Heavy feeling, Bilateral weakness of upper limbs, Asthenia, Exhaustion delirium, Tired |
| Sore throat | Sore throat symptom, Tender larynx, Acute pharyngitis, Infective pharyngitis, Pharyngitis, Irritation of gums and throat, Painful tonsil, Pharyngolaryngitis, Viral pharyngitis, Pain in throat, Exudative pharyngitis |
| Chest pain or pressure | Dull chest pain, Central crushing chest pain, Chest pain on breathing, Non-cardiac chest pain, Musculoskeletal chest pain, Cardiac chest pain, Chest pain, Chest pain, Pain of sternum, Left sided chest pain, Right sided chest pain, Upper chest pain, Crushing chest pain, Retrosternal pain, Bothered by chest pain in last 4 weeks [Reported.PHQ], Chest pain on exertion, Chest pain at rest, Chest pain, Localized chest pain, Squeezing chest pain, Acute chest pain, Chest wall pain, Atypical chest pain, Chest wall tenderness, Central chest pain, Parasternal pain, Costal margin chest pain, Pleuropericardial chest pain, Radiating chest pain, Ischemic chest pain |
| Impairment of consciousness | Syncope and collapse, Vasovagal syncope, Disturbance of consciousness, Convulsive syncope, O/E - clouded consciousness, O/E - decreased level of consciousness, Loss of consciousness, Delirious, Mentally dull, Unresponsive, Feeling faint, Muzzy headed, Seems in a trance, Being out of body, Mentally vague, Brief loss of consciousness, Lightheadedness, Stupor, Syncope, Semiconscious, Intermittent confusion, Decreased level of consciousness, Witnessed syncope, Prolonged loss of consciousness, Complaining of wooziness, Wooziness, Impairment of mental alertness, Minimally conscious state, Hypotensive syncope, Cardiac syncope, Difficult to rouse, Lack of awareness, Clouded consciousness, Moderate loss of consciousness, Daytime somnolence, Cognitive deficit in attention, Acute confusion, O/E - level of consciousness, O/E - mentally confused, O/E - delirious, O/E - semiconscious, O/E - conscious level fluctuating, Seems in a dream |
| Diarrhea | Diarrhea symptom, Psychogenic diarrhea, Diarrhea and vomiting, symptom, Diarrhea of presumed infectious origin, Severe diarrhea, Acute diarrhea, Diarrhea and vomiting, Nausea, vomiting and diarrhea, Diarrhea, Inflammatory diarrhea, Infectious diarrheal disease, Spurious diarrhea - overflow |
| Impairment of taste or smell | Problem of sense of smell, Things smell different, Unusual smell in nose, Loss of taste, Disorder of smell, C/O - anosmia, C/O - loss of taste sense, Loss of sense of smell, Abnormal unpleasant perception of strong scent, Loss of taste posterior one third of tongue, Parosmia, Sense of smell impaired, Hemiageusia, Loss of taste anterior two thirds of tongue, Sense of smell altered, On examination - taste loss anterior 2/3 tongue, O/E-taste loss post 1/3 tongue, O/E - smell abnormal, O/E - anosmia, Congenital anosmia |
| Cough | Unable to cough voluntarily, Difficulty coughing voluntarily, Unexplained cough, C/O - cough, Coughing ineffective, Sputum retention, Finding related to ability to cough, Finding related to ability to cough voluntarily, Finding related to ability to cough up sputum, Respiratory tract congestion and cough, Paroxysmal cough, Painful cough, Clearing throat - hawking, Unable to cough, Cough when swallowing, Effective cough, Difficulty in coughing up sputum, Does cough up sputum, Does not cough up sputum, Able to cough, Does cough, Does not cough, Difficulty coughing, Croupy cough, Productive cough, Persistent cough, Postviral cough, Nocturnal cough, Early morning cough, Increasing frequency of cough, Spasmodic cough, Decreased coughing, Hacking cough, Episodic dry cough, No cough strength, Cough, Cough at rest, Cough suppression, Dry cough, Postural cough, Brassy cough, Cough on exercise, Productive cough -clear sputum, Productive cough -green sputum, Productive cough-yellow sputum, Morning cough, Evening cough, Nocturnal cough / wheeze, Cough with fever, Cough reflex absent, Barking cough, Unable to cough up sputum, Able to cough up sputum |
| Headache | Migraine, Intractable episodic tension-type headache, Idiopathic partial status epilepticus, Muscular headache, Tension-type headache, Lower half migraine, Basilar artery migraine, refractory, Refractory acute confusional migraine, Ophthalmoplegic migraine, refractory, Refractory migraine, Migraine variants, not intractable, Migraine without aura, not refractory, Refractory migraine variants, Intractable ophthalmic migraine, Transformed migraine, Refractory migraine with aura, Refractory migraine without aura, Acute confusional migraine, Ophthalmic migraine, Hindbrain hernia headache, Thunderclap headache, Nervus intermedius neuralgia, Hemicrania continua, Migraine variant with headache, Ophthalmoplegic migraine, Daily headache, Hemiplegic migraine, Migraine with aura, Basilar migraine, Migraine without aura, Retinal migraine, Migraine with persistent visual aura, Infrequent episodic tension-type headache, Frequent episodic tension-type headache, Hypnic headache, Primary thunderclap headache, Common migraine with status migrainosus, Intractable hemiplegic migraine, Hemiplegic status migrainosus, Intractable hemiplegic status migrainosus, New daily persistent headache, Ophthalmic status migrainosus, Retinal status migrainosus, Intractable retinal migraine, Refractory abdominal migraine, Vascular headache, Anterior ethmoidal nerve syndrome, Tympanic plexus neuralgia syndrome, Status migrainosus with aura, Sick headache, Migraine variants, Complicated migraine, Menopausal headache, Painful blind eye, Headache disorder, Migraine with typical aura, Non-familial hemiplegic migraine, Migraine aura without headache, Status migrainosus, Migraine with ischemic complication, Episodic tension-type headache, Low pressure headache, Idiopathic stabbing headache |
| Nausea or vomiting | Uncontrollable vomiting, Vomiting in infants AND/OR children, Vomiting blood - fresh, Bilious vomit on examination, Vomiting, Nausea, Intermittent vomiting, Acute vomiting, Vomiting symptom, Finding of vomiting, Nausea, vomiting and diarrhea, Increased nausea and vomiting, Vomiting in newborn, Vomiting co-occurrent and due to infectious disease, Projectile vomiting, Hematemesis, Intractable nausea and vomiting, Bilious vomiting, Recurrent vomiting, Persistent vomiting, Vomiting without nausea, Nausea present, Nausea and vomiting |
| Rash | Butterfly rash, O/E - scalp rash, O/E - itchy rash, Macular rash, O/E - rash present, Rash of genitalia, Pityriasis rosea, Pustular rash, Erythematous rash, Rash, Rash of scalp, Rash of mouth, Blanching rash, Non-blanching rash, Generalized rash, Rash of groin, O/E - erythematous rash, C/O: a rash, O/E - mouth rash |
| Body/muscle pain and aches | Knee pain, Iliotibial band friction syndrome of right knee, Myalgia/myositis - multiple, Myalgia/myositis - shoulder, Myalgia/myositis - upper arm, Myalgia/myositis - forearm, Myalgia/myositis - hand, Myalgia/myositis -pelvis/thigh, Myalgia/myositis - lower leg, Myalgia/myositis - ankle/foot, Multiple joint pain, Anterior knee pain, Viral myalgia, Lumbar ache - renal, Shoulder joint pain, Arthralgia of the upper arm, Arthralgia of the pelvic region and thigh, Arthralgia of the ankle and/or foot, Pain in thoracic spine, Pain in lumbar spine, Pain of left hip joint, Pain in left knee, Pain in right hip joint, Pain in right knee, Finger joint painful on movement, Thumb joint painful on movement, Intractable low back pain, Hip joint painful on movement, Knee joint painful on movement, Ankle joint - painful on movement, Foot joint - painful on movement, Subtalar joint painful on movement, Toe joint painful on movement, Postural low back pain, Pain in femur, Pain on passive stretch of joint, Segmental dysfunction, Axis pressure pain, Complaining of backache, Acute back pain with sciatica, Posterior compartment low back pain, Facet joint pain, Painful swelling of joint, Pain in the coccyx, Lumbago co-occurrent with right-side sciatica, Metatarsalgia of left foot, Metatarsalgia of right foot, Rib pain, Cervical facet joint pain, Thoracic facet joint pain, Pain on joint movement, Pain on movement of skeletal muscle, Right sided thoracic back pain, Left sided thoracic back pain, Temporomandibular joint painful on movement, Cervical spine painful on movement, Thoracic spine - painful on movement, Lumbar spine painful on movement, Pain of sternum, Shoulder joint painful on movement, Shoulder joint - painful arc, Elbow joint - painful on movement, Wrist joint painful on movement, Low back pain, Mechanical low back pain, Foot joint pain, Musculoskeletal pain, Muscle tension pain, Myofascial pain with referral, Greater trochanteric pain syndrome, Epidemic cervical myalgia, Shoulder joint painful on external rotation, Polymyalgia, Secondary fibromyalgia, Rubella arthralgia, Chronic low back pain, Acute low back pain, Acute thoracic back pain, Thoracic back pain, Myofascial pain, Lumbar facet joint pain, Abdominal muscle pain, Malignant bone pain, Musculoskeletal chest pain, Pain of right shoulder blade, Pain of left shoulder blade, Pain of right acromioclavicular joint, Pain of left acromioclavicular joint, Pelvic girdle pain, Tarsalgia, Low frequency muscle fatigue, Muscle pain, Sacral back pain, Pain in spine, Hip pain, Joint pain, Herniation of lumbar intervertebral disc with sciatica, Arthralgia of temporomandibular joint, Secondary fibrositis, Pain in left lumbar region, Pain in right lumbar region, Pain in lumbar region on palpation, Muscle fatigue, Vertebral joint pain, Fibrositis, Central muscle fatigue, High frequency muscle fatigue, Localized masticatory muscle soreness, Achillodynia, Forearm joint pain, Intractable back pain, Muscle pain with chewing, Costovertebral angle pain, Pain in the rhomboid muscle, Tibial pain, Left paraspinal back pain, Costal chondral thoracic pain, Infrequent episodic arthralgia of temporomandibular joint, Myalgia of pelvic floor, Joint pain of pelvic region, Pain of right sternoclavicular joint, Pain of left sternoclavicular joint, Joint pain in right hand, Joint pain in left hand, Pain of joint of right foot, Pain of joint of left foot, Backache without recent injury, Pain of left temporomandibular joint, Pain of right temporomandibular joint, Pain in left sacroiliac joint, Pain in right sacroiliac joint, Bilateral foot joint pain, Bilateral hip joint pain, Bilateral ankle joint pain, Bilateral shoulder joint pain, Bilateral elbow joint pain, Acute arthralgia of knee, Lumbosacral nerve root pain, Bilateral knee pain, Bilateral sacroiliac joint pain, Bilateral acromioclavicular joint pain, Bilateral temporomandibular joint pain, Pain of joint of bilateral lower legs, Pain of joint of right lower leg, Pain of joint of left lower leg, Pain of bilateral sternoclavicular joints, Bilateral pain of shoulder blades, Chronic back pain, Bone pain, Exacerbation of backache, Pain of left ankle joint, Pain of right ankle joint, Pain of left shoulder joint, Pain of right shoulder joint, Pain of left elbow joint, Pain of right elbow joint, Low back pain co-occurrent and due to bilateral sciatica, Low back pain co-occurrent with neuralgia of left sciatic nerve, Bilateral pain of joint of hands, Backache, Back pain worse on sneezing, C/O - low back pain, C/O - upper back ache, Costal margin chest pain, Peripheral muscle fatigue, Greater trochanteric pain syndrome of right lower limb, Bilateral greater trochanteric pain syndrome of lower limbs, Greater trochanteric pain syndrome of left lower limb, O/E - joint movement painful, Hand joint pain, Sternoclavicular joint pain, Acromioclavicular joint pain, Elbow joint pain, Distal radioulnar joint pain, Wrist joint pain, Metacarpophalangeal joint pain, Proximal interphalangeal joint of finger pain, Distal interphalangeal joint of finger pain, Sacroiliac joint pain, Tibiofibular joint pain, Ankle joint pain, Subtalar joint pain, Talonavicular joint pain, Interphalangeal joint of toe pain, Lumbago with sciatica, Scapulalgia, Bony pelvic pain, Clavicle pain, Tenalgia |
